# Integrating epidemiological and clinical predictors of SARS-CoV-2 infection in students and school staff in the state of São Paulo

**DOI:** 10.1101/2021.06.21.21259213

**Authors:** Fredi A. Diaz Quijano, José Mário Nunes da Silva, Tatiana Lang D’Agostini, Jéssica Pires de Camargo, Nathalia Cristina Soares Franceschi Landi de Moraes, Ricardo Haddad, Maria Cecília Gomes Pereira, Dimas Tadeu Covas, Regiane A. Cardoso de Paula

## Abstract

**Background:** There is a great deal of uncertainty concerning which contexts would be safe for returning to school and about individual criteria that would reduce contact between the infected and susceptible people in the school setting. Therefore, the purpose of this study was to estimate the prevalence of infection by SARS-CoV-2 in students and school staff; and to identify predictors of infection, including both municipal epidemiological indicators and individual variables reported by the participants.

**Methods:** This was a virological survey carried out among students (over 14 years old) and school staff in São Paulo state, between epidemiological weeks 43 to 49 of the year 2020. A self-administrated questionnaire including sociodemographic and clinical information was applied. Moreover, a nasopharynx swab was performed for virological testing (RT-PCR). We evaluate the relationship of COVID-19 epidemiological indicators of the residence municipality with the odds of SARS-CoV-2 infection. For this, a composite index relating recent mortality and previous incidence (RM/PI) was proposed based on the ratio of deaths recorded in the second and third week counted back to the sum of cases during the previous seven weeks (weeks 4 to 10 counted back). We obtained a multiple model using random-effects logit regression integrating epidemiological indicators and individual variables.

**Results:** In total, 3436 participants were included, residents of 72 municipalities. The overall prevalence of infection was 1.7% (95%CI: 1.3%-2.2%). SARS-CoV-2 infection was independently associated with loss of smell, a history of pulmonary disease, and a recent trip outside the municipality. Moreover, the RM/PI index consistently predicted the SARS-CoV-2 infection (adjusted OR: 1.45; 95%CI 1.02-2.04). Based on these associations, we proposed a classification in four groups with different SARS-Cov-2 infection prevalence (0.54%, 1.27%, 3.8%, and 4.13%).

**Conclusion:** Epidemiological and individual variables allowed classifying groups according to the infection probability in a school population of the state of São Paulo. This classification could help guide the return to classes in situations in which epidemiological control is evident, maintaining basic protection measures and increasing vaccination coverage.

## INTRODUCTION

The COVID-19 pandemic continues to spread globally [1,2]. Initially, the pandemic was addressed through the implementation of aggressive public health measures focused on restricting mobility and ensuring physical distance [3,4]. Most countries, including Brazil, imposed the closure of schools to mitigate transmission. This restriction occurred when the exact role of children in the virus spreading and their vulnerability were not known [5,6].

On April 16, 2020, the United Nations Educational, Scientific and Cultural Organization (UNESCO) estimated that 1.57 billion children and young people in more than 190 countries stopped attending school, i.e., around 90% of the students worldwide. In Brazil, there were more than 52 million students who left without face-to-face classes [7]. In São Paulo (state), classes were suspended on March 23 and affected about 6.8 million students from the state, municipal and private schools [8].

The closure of schools reduces the number of contacts within the population and, therefore, the subsequent transmission [9]. However, this measure can also cause considerable damage to children and their families with significant social and economic impacts, mainly on physical and mental health. Moreover, children from low-income families are likely to be affected more adversely than high-income children due to the loss of school services, such as counseling, psychological services, special education, and nutritional support (school meals) [10–14].

On the other hand, when infected, children generally have a milder disease when compared to adults [15–18]. This relative difference in severity affects the cost-benefit ratio associated with school closings. The gradual return to teaching activities began to be discussed worldwide, including Brazil [10–12,19].

Thus, at the end of the last quarter of 2020, several schools restarted some presential activities in municipalities of the state of São Paulo, which had a relatively low COVID-19 incidence [20]. However, this return was carried out irregularly or suspended due to the worsening of the epidemic [21–23]. Moreover, there has been a lack of evidence-based guidelines to reestablish activities Thus, there is a great deal of uncertainty concerning which contexts would be safe for returning to school and which would be individual criteria that would reduce the risk of contact between the infected and susceptible in the school setting.

The purpose of this study was to estimate the prevalence of SARS-Cov-2 infection in students and school staff (including teachers and other employees of the educational institutions). In addition, we aimed to identify predictors of this infection, including municipal epidemiological indicators and individual variables reported by the participants. We believe that these results may guide the formulation of criteria for opening and closing schools.

## MATERIALS AND METHODS

This was a virological survey carried out among students (over 14 years old) and school staff (including teachers and administrative staff) in São Paulo (state). Schools from the Regional Health Secretariats (DRS) municipalities and three from the Metropolitan Region of São Paulo were included, which returned to classes in October or November 2020.

Participants were invited to participate between 19/10/2020 and 01/12/2020, and only people who attended schools were included. This corresponded to the period between epidemiological weeks 43 to 49 of the year 2020, during which there was an apparent stabilization of the incidence and mortality indicators in the participating municipalities (Figure 1) [21].

**Figure 1.**
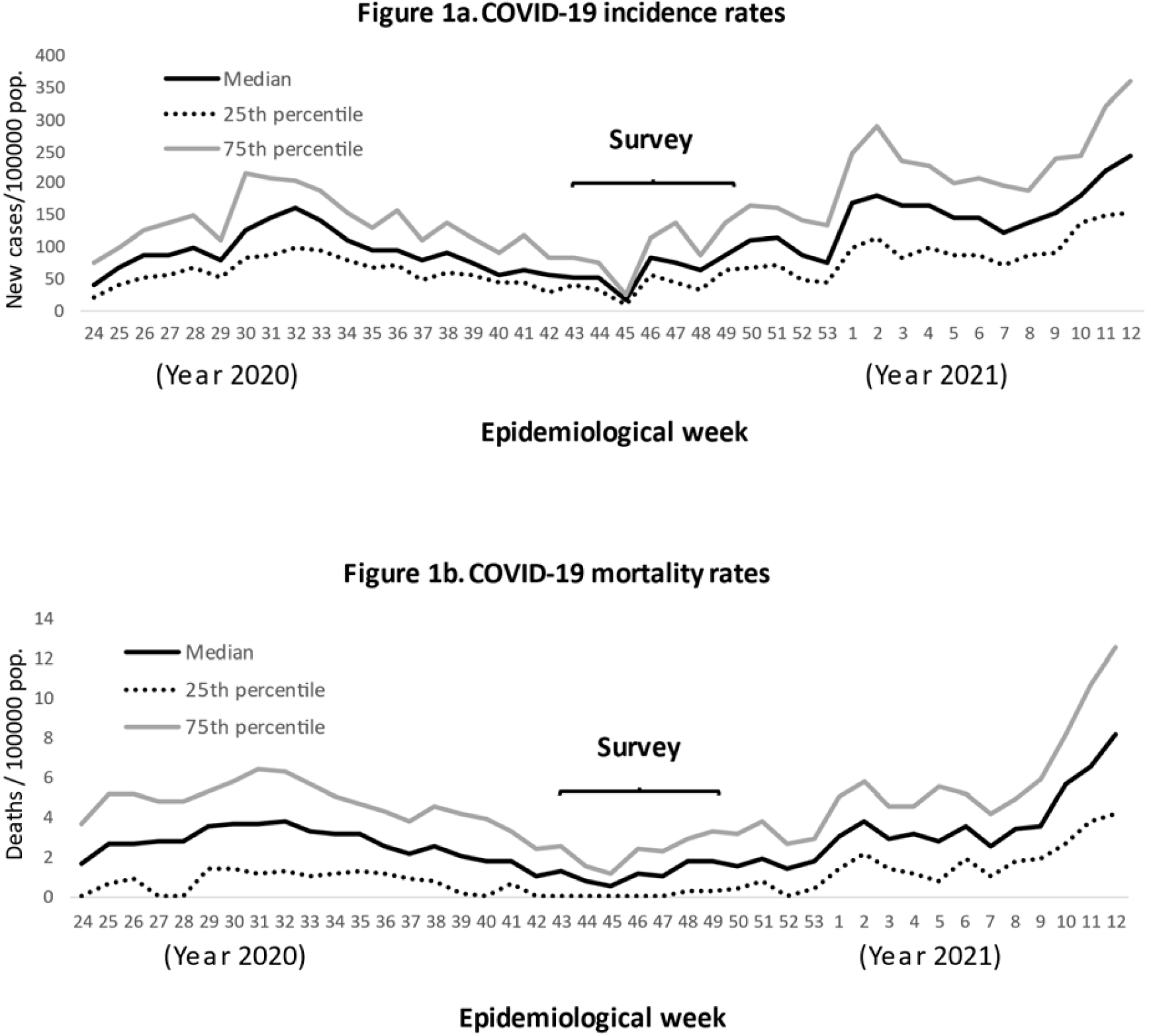
Distribution of weekly COVID-19 incidence and mortality rates in municipalities wherein residing the participants.

After informed consent, each participant was evaluated, including both a questionnaire and a nasopharynx swab for virological testing (RT-PCR). The questionnaires that included demographic and clinical information were self-administered in the school the same day in which of swab sample was collected.

### Diagnosis by laboratory

RNA extraction was performed with the Extracta kit AN viral (Loccus®) in an automated extractor (Extracta 32, Loccus®) following the manufacturer guidelines. SARS-CoV-2 molecular diagnosis was carried out using the kit Gene FinderTM COVID19 Plus RealAmp kit (OSang Healthcare Co. Ltd.) in all laboratories comprising the network which reduces variations related to Ct values.

### Epidemiological indicators

In addition to individual variables collected with the questionnaire, we evaluated whether the epidemiological information of the municipality of residence could predict the state of infection of the participants. The information was obtained through the Foundation System of the State Data Analysis System, SEADE (Fundação Sistema Estadual de Análise de Dados Estatísticos), a national reference center in the production and dissemination of statistics for municipalities and regions of the State of São Paulo [24]. It has released daily newsletters on the situation of COVID-19 in the state based on information provided by the State Department of Health [21].

We first grouped the incidence and mortality data by week, starting every Monday. In this manuscript we will refer to the weeks by listing them backwards, considering the time of the survey as week zero. In that way, the previous one would be week 1, the one before that week number 2, so on. We considered that the epidemiological information may take time to update and what we expected is that the indicators lead to decisions a few days in advance. Therefore, only data from week two and those previous were evaluated as predictors.

We evaluate the relationship of COVID-19 incidence and mortality indicators of the municipality of residence with the odds of infection. We assessed those indicators as event counts, rates and logarithm versions of them. Moreover, observing the weekly values of these indicators in relation to the prevalence by residing municipalities, a composite index was proposed based on the ratio of recent deaths (in the second and third week counted back, considering the survey moment as week zero) to the sum of cases during the previous 7 weeks (weeks 4 to 10 counted back). We named this last indicator as index of recent mortality to previous incidence (RM/PI index).

### Data analysis

Demographic and clinical were entered in an electronic database, and then analyzed using Excel and STATA (version 15.0, Stata Corp LP, College Station, TX). Data analysis included a description of the manifestations potentially attributable to SARSCov2 infection. In a preliminary analysis, the most functional form of the available variables was sought. This included evaluation of the linear relationship between quantitative variables and the frequency of the outcome.

A random-effects logit regression model was used to obtain a multiple model, evaluating both municipality epidemiological indicators and individual characteristics, considering the residing municipality as the clustering variable. A model selection was carried out, evaluating all demographic and clinical variables, as well as epidemiological indicators. After evaluating all the variables, exclusions were made until obtain a model including only terms with p <0.05. With the values predicted by the multiple model obtained, we estimated the area under the ROC curve (AUC). Finally, we proposed an algorithm based on associations to classify groups according to the probability of infection.

### Ethical approval

This study followed Brazilian and International legislation for conducting human research. This research project was approved by the Research Ethics Committee of the School of Public Health of the University of São Paulo (Register number: 4.369.013 and CAAE: 38625120.3.0000.5421).

## RESULTS

In total, 3436 participants aged over 14 years were included, including 1689 students and 1747 school staff members, linked to 84 schools in 16 municipalities. About 85% of participants residing in the same municipality wherein placed the school and the rest were living in neighboring municipalities. Thus, this sample included residents of 72 municipalities.

Among the participants, 60 cases of infection were detected, 30 in students and 30 in school staff members. Therefore, the overall observed prevalence of infection was 1.7% (95% CI: 1.3% - 2.2%). Variables such as age and sex were not associated with the prevalence of infection. On the other hand, we observed that the cases of SARS infection manifested a significantly higher frequency of fever, cough, loss of smell, a history of lung disease and recent trip outside the municipality (Table 1).

**Table 1.** Comparative description of participants according to SARS-CoV-2 infection.

| **Variable** | **Total (n=3436)** | **SARS-PCR**<br>**positive (n=60)** | **SARS-PCR**<br>**negative (n=3376)** | **p-value** |
| --- | --- | --- | --- | --- |
| Age (years)–median (IQR) | 26.3 (16.9 – 47.3) | 21 (17.1 – 44.6) | 26.5 (16.9 – 47.4) | 0.94 |
| Sex Male – No. (%) | 1310 (38.1%) | 29 (48.3%) | 1281 (37.9%) | 0.11 |
| Student | 1689 (49.2%) | 30 (50%) | 1659 (49.1%) | 0.90 |
| Symptoms |  |  |  |  |
| Fever | 46 (1.3%) | 3 (5%) | 43 (1.3%) | 0.05 |
| Myalgia | 478 (13.9%) | 10 (16.7%) | 468 (13.9%) | 0.57 |
| Chills | 84 (2.4%) | 0 (0%) | 84 (2.5%) | 0.40 |
| Coryza | 474 (13.8%) | 9 (15%) | 465 (13.8%) | 0.71 |
| Cough | 292 (8.5%) | 11 (18.3%) | 281 (8.3%) | 0.02 |
| dor_gargan~p | 303 (8.8%) | 5 (8.3%) | 298 (8.8%) | 1 |
| loss of smell | 54 (1.6%) | 4 (6.7%) | 50 (1.5%) | 0.01 |
| loss of taste | 46 (1.3%) | 1 (1.7%) | 45 (1.3%) | 0.56 |
| vomit | 47 (1.4%) | 1 (1.7%) | 46 (1.4%) | 0.57 |
| Palpitation | 119 (3.5%) | 1 (1.7%) | 118 (3.5%) | 0.72 |
| Diarrhea | 157 (4.6%) | 1 (1.7%) | 156 (4.6%) | 0.52 |
| Breathing difficulty | 144 (4.2%) | 4 (6.7%) | 140 (4.1%) | 0.32 |
| Clinical history |  |  |  |  |
| Cardiac disease | 45 (1.3%) | 1 (1.7%) | 44 (1.3%) | 0.55 |
| HIV infection | 3 (0.1%) | 0 (0%) | 3 (0.1%) | 1 |
| Renal disease | 46 (1.3%) | 0 (0%) | 46 (1.4%) | 1 |
| Liver disease | 26 (0.8%) | 1 (1.7%) | 25 (0.7%) | 0.37 |
| Pulmonary disease | 133 (3.87%) | 7 (11.7%) | 126 (3.73%) | 0.008 |
| Hypertension | 598 (17.4%) | 10 (16.7%) | 588 (17.4%) | 1 |
| Diabetes | 168 (4.9%) | 2 (3.3%) | 166 (4.9%) | 1 |
| Exposure background |  |  |  |  |
| Contact with a confirmed COVID-19 case | 563 (16.4%) | 12 (20%) | 551 (16.3%) | 0.48 |
| Contact with a suspected COVID-19 case | 94 (2.7%) | 2 (3.3%) | 92 (2.7%) | 0.68 |
| Exposed to hospital environment | 624 (18.2%) | 13 (21.7%) | 611 (18.1%) | 0.50 |
| Recent trip outside of municipality | 458 (13.3%) | 15 (25%) | 443 (13.1%) | 0.01 |

Different versions of the incidence or mortality indicators were not associated with the infection prevalence (Table 2). However, the RM/PI index consistently predicted the SARS-CoV-2 infection (Figure 1). Thus, each increase in one percentage unit in the index was associated with a 49% increase in the odds of SARS-CoV-2 infection (OR: 1.49; 95%CI 1.06 – 2.08).

**Table 2.** Evaluation of municipal epidemiological indicators to predict the individual status of SARS-CoV-2 infection.

| Epidemiological indicator | Formula* | OR (95% CI) † | p-value |
| --- | --- | --- | --- |
| Accumulated new cases | $c_{2-\infty}$ | 1 (1-1) | 0.14 |
| Accumulated deaths | $d_{2-\infty}$ | 1.01 (1 – 1.02) | 0.12 |
| Logarithmic version of accumulated cases | $\ln(1 + c_{2-\infty})$ | 0.96 (0.75 – 1.23) | 0.73 |
| Logarithmic version of accumulated deaths | $\ln(1 + d_{2-\infty})$ | 0.98 (0.71 – 1.34) | 0.88 |
| Cumulative incidence rate | $\frac{c_{2-\infty}}{p} 10^5$ | 1 (1 – 1) | 0.76 |
| Cumulative mortality rate | $\frac{d_{2-\infty}}{p} 10^5$ | 0.89 (0.62 – 1.27) | 0.52 |
| Logarithmic version of cumulative incidence rate | $\ln(\frac{1 + c_{2-\infty}}{p})$ | 0.86 (0.44 – 1.7) | 0.67 |
| Logarithmic version of cumulative mortality rate | $\ln(\frac{1 + d_{2-\infty}}{p})$ | 1.05 (0.58 – 1.9) | 0.88 |
| Index of recent mortality to previous incidence (RM/PI) | $\frac{d_{2-3}}{c_{4-10}} 10^2$ | 1.49 (1.06 – 2.08) | 0.02 |

\* Formulas to define the epidemiological indicators, being, “ $c_i$ ” the number of new COVID-19 cases in the $i^{th}$ week counted backward starting in the examen week (zero), “ $c_{i-j}$ ” the summatory of new cases from the week $i^{th}$ to week $j^{th}$ ; $d_i$ and $d_{i-j}$ , the number of COVID-19 deaths in the same periods; “ $p$ ” the population estimated for the municipality; ∞ refers to first week of pandemic or the last week of the backward counting; and $\ln$ the natural logarithm, consider the epidemiological indicators presented in the table 1 and their associations with the prevalence odds ratio calculated in a random effect logit model.
† Odds Ratio (OR) regarding the associations between the epidemiological indicator and the SARS-CoV-2 infection prevalence, calculated in a random-effect logit model considering the residing municipality as the clustering variable.

**Table 3.** Multiple model to predict SARS-CoV-2 infection.

| Predictor | OR (95% CI) | p-value |
| --- | --- | --- |
| Index of RM/PI | 1.45 (1.02 - 2.04) | 0.04 |
| Pulmonar disease | 3.56 (1.56 - 8.11) | 0.003 |
| Loss of smell | 5.12 (1.74 - 15.05) | 0.003 |
| Recent trip | 2.13 (1.16 - 3.93) | 0.02 |

In the multiple model, SARS-Cov-2 infection was independently associated with the municipal RM/PI index and the following individual variables: loss of smell, pulmonary disease and history of recent trip (Table 2). The resulting model exhibited an area under the ROC curve of 65.7% (95%CI: 58.6% - 72.9%).

Subsequently, based on these associations, we proposed and algorithm to predict infection by integrating both epidemiological indicators and individual variables associated with SARS-CoV-2 infection (Figure 2). This algorithm leads to four categories of risk:

**Figure 2.**
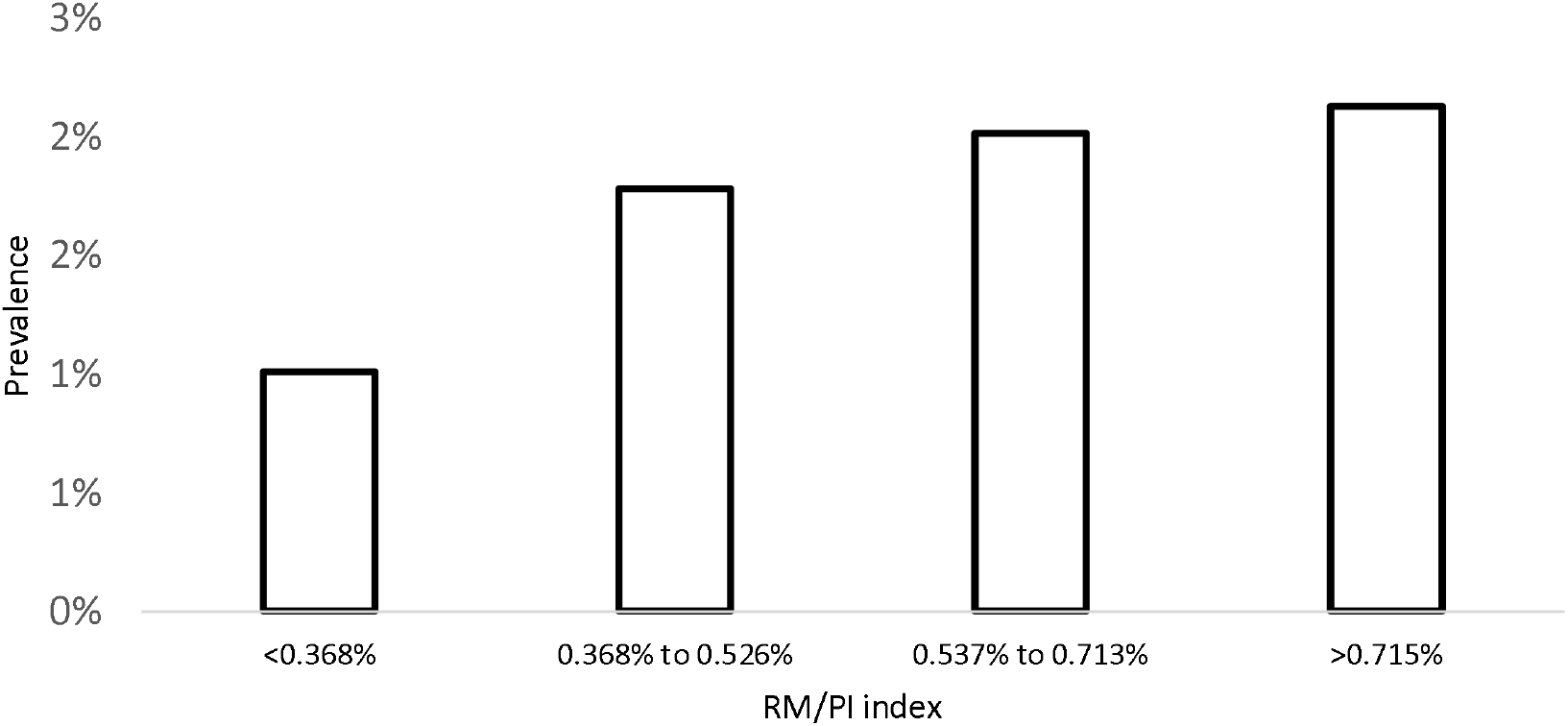
Prevalence of SARS-CoV-2 infection according index of RM/PI.

- Group 1: People without any individual characteristics associated with SARS-CoV-2 (loss of smell, pulmonary disease or history of recent trip), who resided in municipalities with an index lower than 0.4%.
- Group 2: People without any of individual characteristics associated with SARS-CoV-2 residing in municipalities with an index between 0.4% and 1%.
- Group 3: People with one or more characteristics associated with SARS-CoV-2 residing in municipalities with an index between 0.4% and 1%.
- Group 4: People residing in municipalities with an index higher than 1% (independent of individual characteristics).

These groups exhibited a SARS-Cov-2 infection prevalence of 0.54%, 1.27%, 3.8% and 4.13%, respectively (Figure 3). This classification exhibited an ROC curve area similar to that obtained with the multiple model (67.7%; 95%CI: 61.3% – 74%).

**Figure 3.**
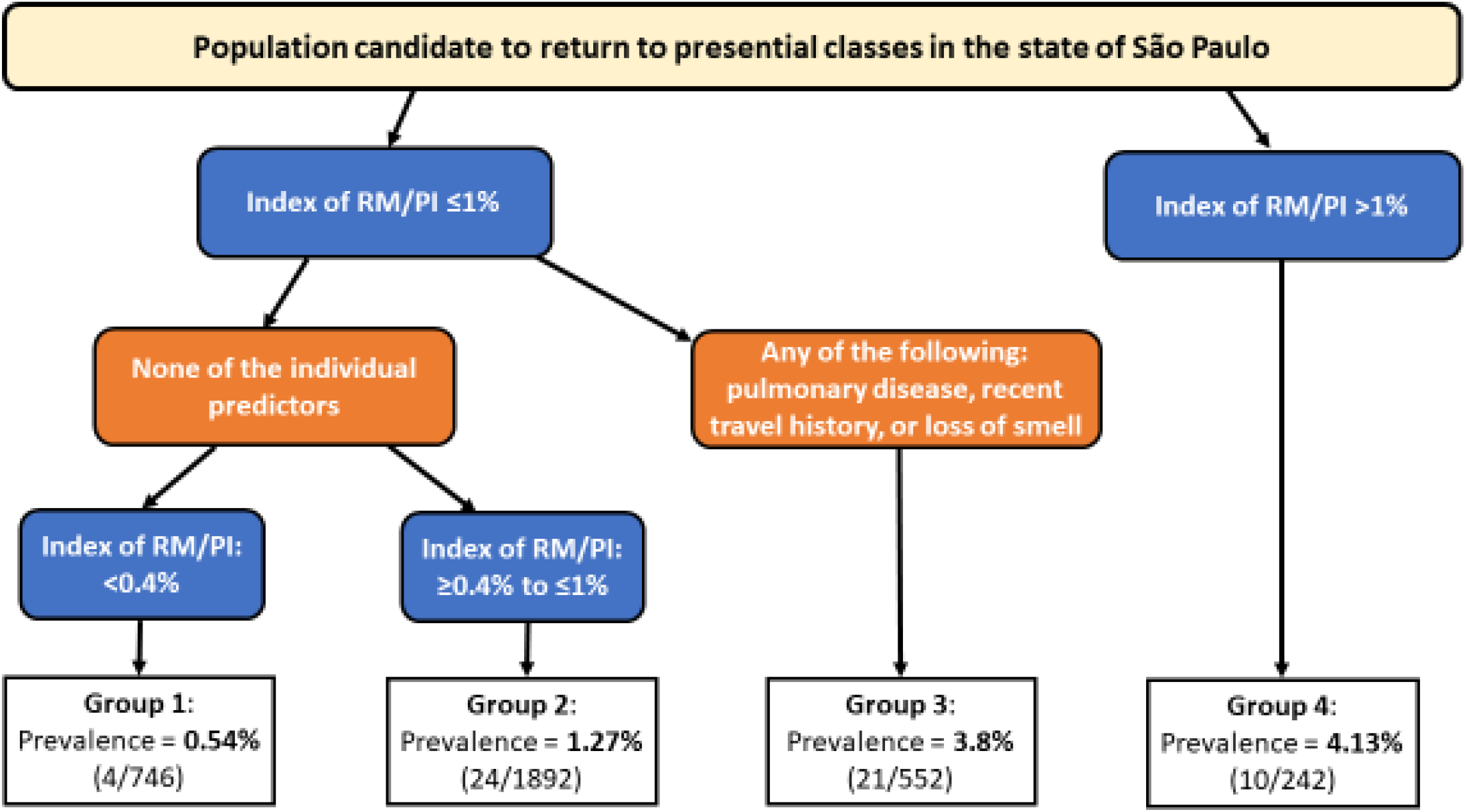
Algorithm to classify the school population according to epidemiological and clinical variables associated with SARS-CoV-2 infection prevalence.

## DISCUSSION

In this work, an estimate of the prevalence of SARS-CoV-2 infection among the school population of the state of São Paulo was carried out. This prevalence should be interpreted considering that the included participants were those who attended face-to-face classes during an epidemiological context of apparent stability in relation to incidence and mortality. In this context, we identified a predictive model that integrated both epidemiological indicators and individual variables easily assessable through a questionnaire.

Our multiple model included an index relating recent mortality and previous incidence. This index does not consider the size of the municipality. For this reason, we evaluated the municipal population but it was neither associated with the prevalence of the infection nor did it affect the other associations (data not shown). Moreover, we observed that neither incidence and mortality measures separately had a direct association with the prevalence recorded in our survey.

The use of incidence and mortality indicators to make individual predictions can be contentious. Among other things, because different aspects such as underreporting and temporal trends must be considered. In this sense, the RM/PI index may incorporate these aspects. For example, when referring to a recent indicator (such as mortality) in relation to a previous one (of incidence), this index would increase when incidence tends to grow (assuming that the case fatality rate remains as a constant or that at least does not decrease). On the other hand, considering that underreporting tends to be higher in mild cases, a lower sensitivity of the surveillance system would affect the denominator (the recorded incidence) of the index more than the numerator. Therefore, it is expected that the proposed RM/PI index will increase when either of an increasing trend of actual cases, an elevated underreporting or both occur. In our survey, this simple index showed a gradient with the prevalence of SARS-Cov-2 infection among the participants, and this association remained consistent in the multiple model.

Besides this municipal epidemiological indicator, we identified easily measurable individual predictors. Among these, the manifestation of pulmonary disease was associated with infection. This could be explained by different mechanisms. For example, the referred disease could refer to COVID-19 itself. On the other hand, antecedents of pulmonary problems could predispose to acquisition and permanence of infections [25,26].

Additionally, the loss of smell showed a strong association with the odds of infection, which is consistent with clinical studies that report a significant frequency of this manifestation in patients with COVID-19 [27–29]. Finally, a recent trip outside the municipality was also an antecedent associated with the infection. This association highlights the importance of mobility as a risk factor for contagion [30–33].

The associations identified, particularly the epidemiological ones, must be interpreted considering the context. Our model was obtained in a situation that suggested an apparent epidemic control. Therefore, it could be more applicable if the levels of morbidity and mortality are relatively stable or decreasing. In this sense, the proposed algorithm could guide the order of return to the classes, starting with municipalities with very low RM/PI index and only for people without clinical predictors of infection. However, because the infection probability was not zero in any predictable circumstance, protective measures such as masks and distancing should always be maintained [34]. Moreover, all municipalities must prioritize the goal of high vaccination coverage [35].

In conclusion, we estimated the prevalence of active SARS-Cov-2 infection in the school population of municipalities of the state of São Paulo. Although the observed prevalence was relatively low, epidemiological and individual variables allowed classifying groups according to the infection probability. We consider that this classification could help guide the return to classes in situations in which epidemiological control is evident, maintaining basic protection measures and increasing vaccination coverage.

## Data Availability

The data that support the findings of this study are available on request from the corresponding author.

## Authors’ contributions

FADQ conceived the study, participated in its design and coordination, carried out the data analysis and prepared the first draft of the manuscript. JMNS Participated in its design, revised the manuscript, and approved the final version of the manuscript for publication. TLDA, JPC, NCSFLM, MCGP and RACP participated in the conception, design and coordination of the study. DTC and RH conceived and designed the flow of samples in the diagnostic area. All authors provided relevant input for the writing, carried out reviews, and read and approved the final manuscript.

Thus, each author participated sufficiently in the work to take public responsibility for appropriate portions of the content and, therefore, agreed to be accountable for all aspects in ensuring that questions related to the accuracy or integrity of any part of the work are appropriately investigated and resolved.

## Acknowledgments

The authors would like to thank the Butantan Foundation for the financial support for the inputs, reagents and technical team of the Strategic Laboratory of the Diagnosis and Quality Control Center.

